# Dissecting antibody responses to cardiac receptors in patients with myocardial infarction

**DOI:** 10.1101/2025.11.03.25339371

**Authors:** Johanna Siegel, Daniel Beer, Stasa Janjatovic, Anne Auer, Lea-Sophie Zipp, Evelyn Kozuch, Tobias Krammer, Marie Bielenberg, Antoine-Emmanuel Saliba, Clément Cochain, Stefan Störk, Peter Heuschmann, Caroline Morbach, Niklas Beyersdorf, Matthias Nahrendorf, Viacheslav O. Nikolaev, Ulrich Hofmann, Stefan Frantz, Hedda Wardemann, Margarete Heinrichs, Katrin G. Heinze, Katrin Streckfuss-Bömeke, DiyaaElDin Ashour, Gustavo Campos Ramos

**Affiliations:** Department of Internal Medicine I, University Hospital Würzburg, Würzburg, Germany; Comprehensive Heart Failure Center, University Hospital Würzburg, Würzburg, Germany; Institute of Pharmacology and Toxicology, Julius-Maximilians-Universität Würzburg (JMU), Würzburg, Germany; Rudolf Virchow Center for Integrative and Translational Bioimaging, Julius-Maximilians-Universität Würzburg (JMU), Würzburg, Germany; Clinic for Cardiology and Pneumology, Georg-August University Göttingen and DZHK (German Center for Cardiovascular Research), Partner Site Göttingen, Germany; Helmholtz Institute for RNA-based Infection Research (HIRI), Helmholtz-Center for Infection Research (HZI), Würzburg, Germany; Institute of Experimental Cardiovascular Research, University Medical Center Hamburg-Eppendorf, Hamburg, Germany; Institute of Experimental Biomedicine, University Hospital Würzburg, Würzburg, Germany; Paris Cardiovascular Research Center, Université Paris Cité, INSERM U970, Paris, France; Dept. Clinical Research & Epidemiology, Comprehensive Heart Failure Center, University Hospital Würzburg and Dept. Internal Medicine, University Hospital Würzburg, Würzburg, Germany; Institute of Clinical Epidemiology and Biometry, Julius-Maximilians-Universität Würzburg (JMU), Würzburg, Germany; Clinical Trial Center, University Hospital and Julius-Maximilians-Universität Würzburg (JMU), Würzburg, Germany; Institute for Virology and Immunobiology, Julius-Maximilians-Universität Würzburg (JMU), Würzburg, Germany; Center for Systems Biology, Massachusetts General Hospital and Harvard Medical School, Boston, MA, USA; Department of Radiology, Massachusetts General Hospital and Harvard Medical School, Boston, MA, USA; Division of B Cell Immunology, German Cancer Research Center, Heidelberg, Germany; Medical Clinic I, Cardiology and Angiology, Justus-Liebig-University, Giessen, Germany

**Keywords:** Myocardial Infarction, Antibodies, B cells, BCR, Adrenergic Receptors

## Abstract

Myocardial infarction (MI) triggers the production of heart-reactive serum antibodies, but their antigen specificities and impact on cardiomyocyte function remain poorly understood. Using single-cell RNA and B cell receptor sequencing (scRNA/BCRseq) on plasmablasts isolated from patients with MI combined with antibody engineering, we identified B cell clones expanded in response to MI, and generated a panel of 17 recombinant monoclonal antibodies having the same binding domains as those identified in patients scBCR datasets (MI-mAb). This approach enabled us to map antibody specificities and dissect their functional impact on cardiomyocytes. Our findings reveal that post-MI B cell responses target cardiac receptors, including extracellular epitopes mapped to beta adrenergic receptors (β-ARs). Notably, some of the β-AR-specific MI-mAbs induced intracellular cAMP signaling and ultimately modulated cardiomyocyte excitation-contraction coupling. Taken together, these observations provide mechanistic evidence for expanding B cells in the context of MI, with the production of antibodies targeting cardiac receptors that can impact myocardial pathophysiology.

## INTRODUCTION

Myocardial infarction (MI) and heart failure (HF) can trigger adaptive immune responses directed against cardiac antigens released by dying cardiomyocytes ^1–4^. Elevated serum titers of antibodies targeting cardiac sarcomere elements and cardiac receptors have been reported in mouse models of cardiac injury and in various clinical cohorts ^5–8^. A previous study reported that MI patients with positive serology for troponin-specific antibodies had a reduced ejection fraction ^9^. Moreover, a recent retrospective study monitoring HF patients hospitalized for acute cardiac decompensation found that patients who newly developed heart-reactive antibodies exhibited a higher risk of death and rehospitalization compared to patients who did not produce cardiac antibodies ^10^.

Whether heart-reactive antibodies are merely biomarkers of cardiac injury or actively contribute to disease progression remains unresolved. However, evidence from experimental models supports the latter. Independent laboratories have reported that immunoglobulin-deficient mice subjected to experimental MI demonstrated preserved systolic function and blunted adverse cardiac remodeling compared to infarcted WT controls ^11,12^. Further, Zouggari and colleagues reported that infarcted mice treated with a B cell-depleting anti-CD20 antibody showed reduced myocardial inflammation and improved functional outcomes, and similar B cell-depletion is now being tested in MI patients ^13^. These observations indicate that B cells and antibodies promote post-MI adverse cardiac remodeling and fuel the development of ischemic HF. Nevertheless, the relevant antigen specificities and the mechanisms by which heart-reactive antibodies impact myocardial function remain elusive.

To bridge this knowledge gap, we combined single-cell RNA and B cell receptor (BCR) sequencing (scRNA/BCRseq) and antibody engineering technologies to track antigen-specific B cell responses in patients with MI. We produced a panel of monoclonal antibodies encoding the antigen-binding variable regions identified in silico, effectively recapitulating those produced in vivo (MI-mAb). This approach revealed that post-MI antibody responses target receptors relevant to cardiac function, including β adrenergic receptors (β-ARs), thus enabling us to dissect how myocardial antibodies affect cardiomyocyte excitation and contraction. By expanding the field beyond the constraints of classical serological approaches, our study provides in-depth cardiac epitope mapping and a deeper understanding of how antibodies binding cardiac receptors contribute to MI pathophysiology. Additionally, by implementing a pipeline to rapidly produce and characterize antibodies from individual patients, our study paves the way for monitoring immune mechanisms in myocardial diseases in a patient-specific manner.

## RESULTS

### MI induces the rapid mobilization of clonally expanded plasmablasts in humans

To investigate B cell clonal dynamics and antibody production following MI, ten patients diagnosed with a first acute ST-elevation MI (STEMI) were recruited for a longitudinal characterization and serial sampling at index hospitalization and follow-ups on day 5, month 4 and month 12 post-MI. (Figure 1A; Table S1). As controls, we analyzed samples from age- and sex-matched healthy subjects previously recruited as part of a larger population-based study in Würzburg ^14^. To investigate the presence of heart-reactive antibodies in the plasma of these subjects we performed indirect immunofluorescence staining on human induced-pluripotent stem cell-derived cardiomyocytes (iPSC-CM) ^15^. Prior to each experiment, ventricular iPSC-CM quality was assessed to confirm high purity (80-99%) and successful differentiation of iPSC-CM harbouring regular Z-disc (α-actinin) and M-line (titin exons 8/9) sarcomeric structures and expressing the ventricular marker *MLC2v* (Fig. S1A-C). The viability of cells treated with Ctrl-IgG and MI-mAbs was determined by the Resazurin assay (Fig. S1D). Plasma obtained from MI-patients revealed heterogeneous antibody responses against human iPSC-CMs, with some patients exhibiting plasma IgGs binding to iPSC-CM sarcomere structures (P5) and (peri)nuclear structures (P7, P8, P12), while others showed no specific binding (Fig. 1B). Notably, all MI patients with humoral immunity to iPSC-CMs presented cardiomyocyte-specific IgGs already at index hospitalization (Fig. 1C).

**Fig. 1:**
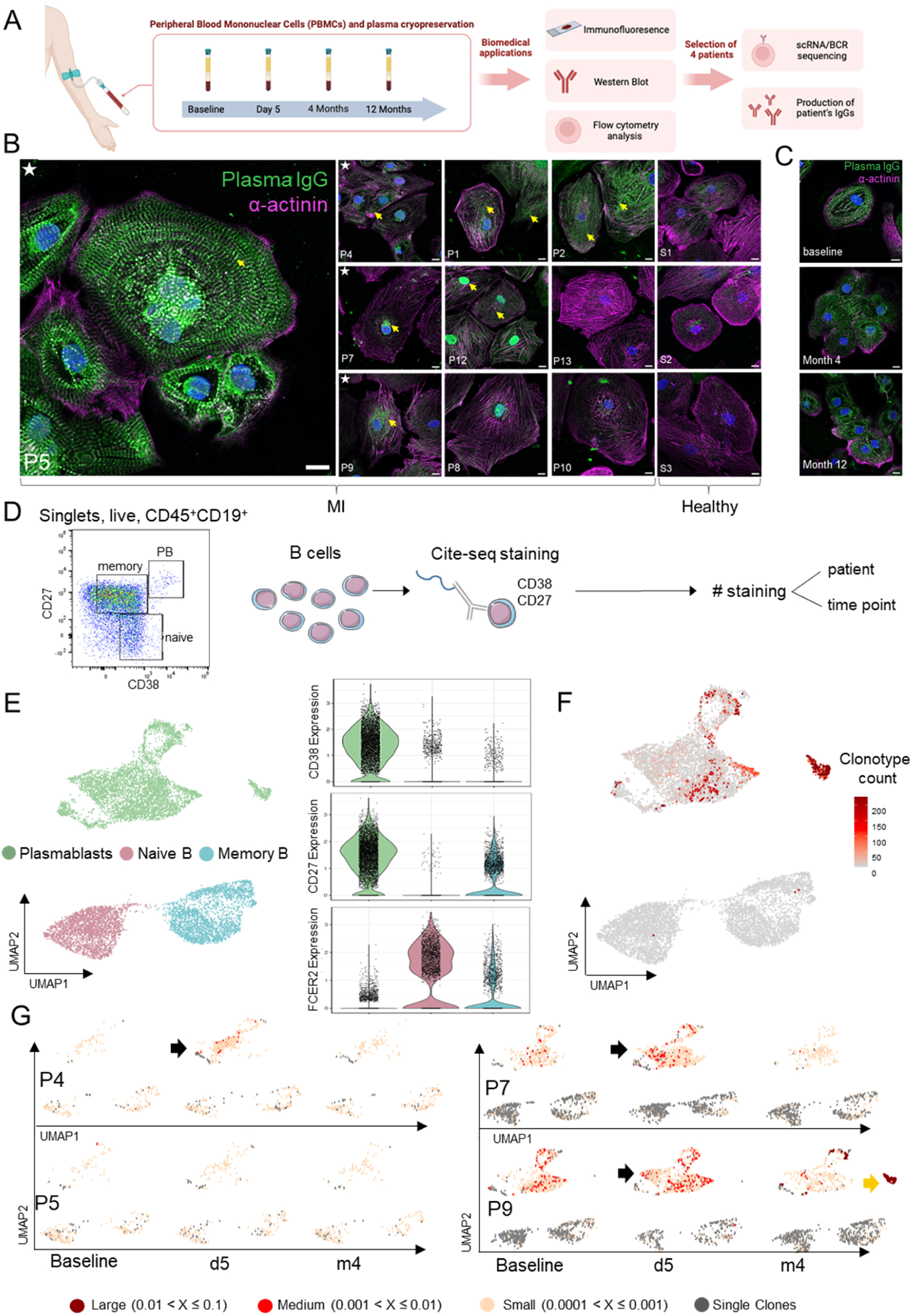
MI induces a rapid mobilization of clonally expanded antibody-secreting cells in patients. **(A)** Study design. **(B)** Immunofluorescence imaging using iPSC-CMs incubated with patient plasma reveals the presence of myocardial antibodies in some MI patients (P), but not in healthy subjects (S1 - S3). Yellow arrows indicate plasma IgG binding to iPSC-CMs. White stars indicate patients selected for scRNA/BCRseq. **(C)** Time-course analysis at index hospitalization (baseline) (a), 4 months (b), and 12 months (c) post-MI in a representative patient. Green: plasma IgG; magenta: α-actinin; blue: Hoechst. Scale bar: 10 µm. **(D)** Cell purification strategy. Peripheral blood B cells from each patient were defined as naïve (CD19^+^CD27^-^CD38^int^), memory (CD19^+^CD27^+^CD38^-/+^), and plasmablasts (CD19^+^CD27^high^CD38^high^), plus schematic representation of CITE-seq and TotalSeq-C hashtag antibody staining strategy. **(E)** UMAP plot representation of 10,225 B cells clustered into naïve B cells, memory B cells, and plasmablasts. Data aggregated from four different patients and all time points (left). Violin plots representing the expression of marker genes (*CD38, CD27, FCER2*) for cluster identification (right). **(F)** Feature plot of B cell receptor sequencing analyses revealing clonal size distribution across the B cell clusters. Color intensity indicates clonotype count, and the higher the clonotype count, the more cells express the same B cell receptor. **(G)** UMAP plots of B cell receptor sequencing analyses depicting clonal size distribution in each patient over time.

Based on these observations, we selected four patients showing plasma antibody reactivity to cardiomyocytes (P4, P5, P7, P9, indicated by ★ in Figure 1B), for more detailed characterization of the BCR clonal dynamics among circulating plasmablasts, which likely reflect the MI-induced acute B cell response. Thus, we purified different B cell subsets from PBMC samples, and post-sorting enriched for plasmablasts (CD45^+^CD19^+^CD27^high^ CD38^high^). Additionally, memory B cells (CD45^+^CD19^+^CD27^+^CD38^-/+^) and naïve B cells (CD45^+^CD19^+^CD27^-^CD38^int^) were purified and served as filler, to allow for efficient sorting and sequencing of rare plasmablasts, and as control cells (Fig. S2A, Fig. S3). Cells were pooled at a final ratio of 1:1:2 (naïve : memory : plasmablasts). We then performed scRNA/BCRseq at the time of index hospitalization (baseline), five days and four months after MI, using barcoded hashtag antibodies for later demultiplexing (Fig. 1D). Hashtag antibodies were used to identify each individual patient and timepoint, whereas CITE-seq antibodies for CD27 and CD38 were utilized to refine downstream identification of these clusters. After quality control (Fig. S2B-C), 10,225 B cells were analyzed and annotated to three distinct B cell subset clusters based on differential gene expression and CITE-seq profiles: plasmablasts (*PRDM1, TNFRSF17, CD38, IGHG1, MKI67, DERL3*), naïve B cells (*TCL1A, BACH2, AFF3, FCER2, IL4R*), and memory B cells (*GPR183, SCIMP, CD82*) (Fig. 1E, Fig. S2D). BCR clonotype analysis revealed the highest degree of clonal expansion among plasmablasts (Fig. 1F) and at day 5 post-MI in P4, P7, and P9 (Fig. 1G). Moreover, P9 showed a unique cluster of highly expanded cells emerging exclusively four months post-MI. Taken together, these findings highlight a rapid development of clonally expanded plasmablasts in MI patients.

### Recombinant MI-mAbs as tools for dissecting patient-specific immune mechanisms in MI

The expanded plasmablast lineages expressed different immunoglobulin isotypes (IgM, IgG1, IgG2, IgG3, and IgA) and varied in their mutation levels. To functionally characterize the antibodies produced by these cells, we cloned their immunoglobulin (Ig) heavy and light chain variable region genes into expression vectors containing the human Igγ1 and Igκ1 constant regions to produce recombinant monoclonal antibodies with the same antigen specificity as those arising in the B cell from which the antibody was derived ^16^ (Fig. 2A). In total, we generated 17 mAbs selected from different patients (six from P4, one from P7, and 10 from P9) based on their high degree of clonal expansion and/or mutation rates (Table S2). First, to assess whether the individual MI-mAbs show binding to cardiomyocytes, we preformed indirect immunofluorescence imaging using iPSC-CM, as described in Figure 1. As shown in Fig. 2B, an initial screening revealed that 6/17 patient-derived recombinant mAbs derived from P4 and P9 showed strong binding to iPSC-CMs.

**Fig. 2:**
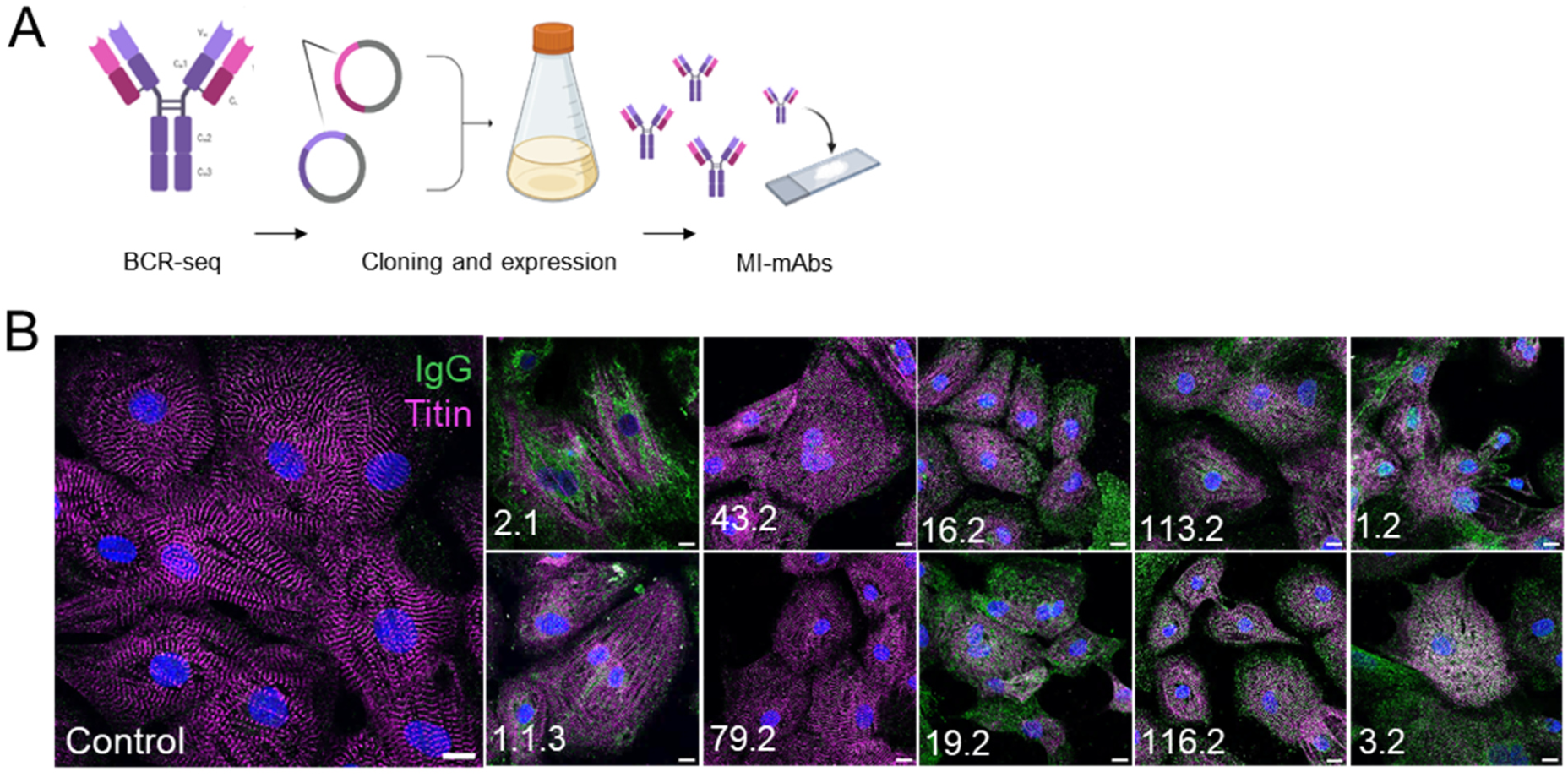
MI-mAbs as tools for dissecting patient-specific immune mechanisms in MI. **(A)** Schematic representation of mAb expression. **(B)** Immunofluorescence images of various patient-derived MI-mAbs (and isotype control IgG) incubated with human iPSC-CMs. IgG concentrations and laser power were adjusted based on binding affinities of MI-mAbs: 2.1 and 1.1.3 0.5% 10 µg/ml, control, 43.2, 79.2, 16.2, 19.2, 113.2, 1.2, and 3.2 1% 10 µg/ml, 116.2 1% 1µg/ml. Green: patient-derived MI-mAbs; magenta: titin; blue: Hoechst. Scale bar: 10 µm.

### MI-mAbs target epitopes on the first and second ß-ARs extracellular loops

Next, we sought to uncover antigens targeted by the MI-mAbs generated in our study. In light of previous studies reporting that HF patients can present antibodies targeting β-adrenergic receptors (β-ARs) ^6–8,17–19^, we decided to focus on possible antibody binding to cardiac receptors expressed on the cardiomyocyte surface. Thus, we designed a panel of peptides covering the β1- and β2-AR extracellular domains and determined the reactivity of mAbs to this peptide library by enzyme-linked immunosorbent assay (ELISA). This approach confirmed that some mAbs bound to peptides derived from the β1-/β2-ARs extracellular domains, suggesting cross-reactivity with both receptors (2.1, 16.2, 19.2, 79.2, 116.2, 1.1.3; Fig. 3A). It is worth noting that recombinant mAbs from different single B cells belonging to clonotype 1.1 differed in their somatic mutation profiles and CDR3 sequences (Fig. 3B). In a previous study we reported the identification of β1-AR-derived epitopes presented to CD4^+^ T helper (Th) cells on HLA-DRB1*13 in MI patients ^20,21^. Thus, the observations of converging T and B cell responses targeting β-AR in patients raises the possibility of a B-Th crosstalk favoring the production of cardiomyocyte-reactive antibodies ^22^.

**Fig. 3:**
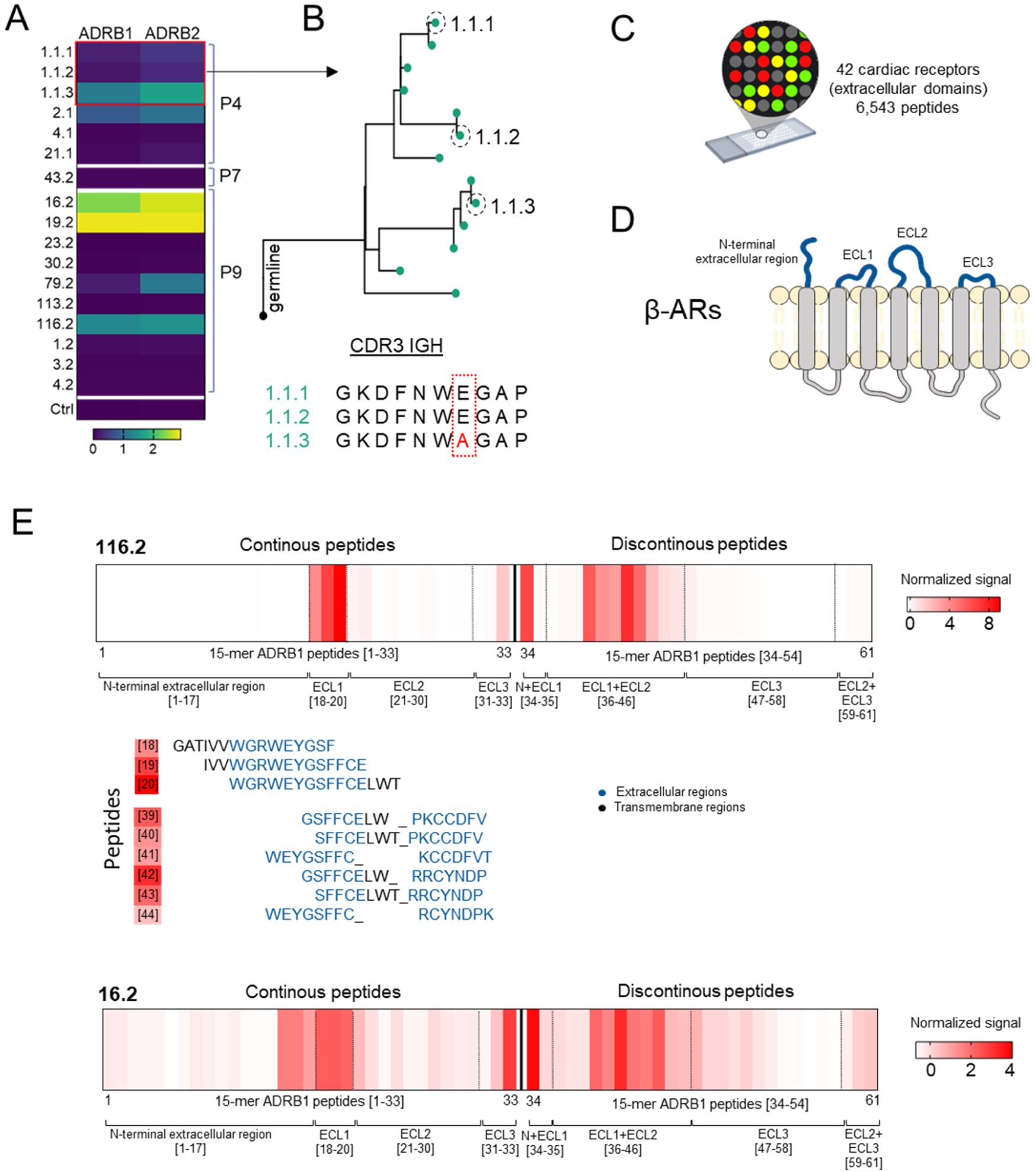
ADRB1 epitope mapping based on antibody array assay. **(A)** Heat map visualizing MI-mAbs and control binding to extracellular peptides derived from ADRB1 and ADRB2, as measured by indirect sandwich-ELISA. The colors indicate respective OD values with yellow representing the highest binding. **(B)** Phylogenetic tree demonstrating the evolutionary trajectory of the B cell merged heavy and light chain clones identified under lineage 1.1. The dashed circle highlights the BCRs (1.1.1, 1.1.2, 1.1.3) chosen for cloning (top). Comparison of complementary-determining region 3 (CDR3) sequences within IGHG2, with mutations highlighted inside the red rectangle (bottom). **(C)** Design of the peptide array including extracellular regions from 42 cardiac receptors with a total of 6,543 peptides. **(D)** Schematic representation of ß-ARs, highlighting its extracellular domains (N-terminus, extracellular loops 1 – 3; ECL1, ECL2, ECL3). **(E)** Epitope mapping for MI-mAbs 116.2 and 16.2. Overlapping peptides with a normalized signal ≥ 3 were considered as high confidence epitopes.

Next, to map the exact epitopes targeted by MI-mAbs, we designed a custom peptide array consisting of 6,543 overlapping peptides spanning the extracellular domains of 42 cardiac membrane proteins, including the β-ARs (Fig. 3C, Table S3). The array included continuous and discontinuous (representing conformational epitopes) 15-mer peptides, against which the MI-mAbs 2.1, 1.1.3, 16.2, 19.2, 79.2, 116.2, 1.2, 3.2, and 43.2 were tested. The β-AR peptide panel covered the N-terminal extracellular region and extracellular loops 1 - 3 (ECL1 – 3, Fig. 3D). Mapping of the binding of mAbs 116.2 and 16.2 showed high-confidence binding to overlapping linear peptide fragments mapping to the β1-AR ECL1, as well as to discontinuous conformational epitopes spanning the β1-AR ECL1 and ECL2 regions (Fig. 3E). Although the normalized signal intensities varied between the two mAbs, the identified epitopes were similar. It should be noted that the signal intensity in the peptide array reflects relative binding strength and does not directly correspond to binding affinity.

### β-AR antibodies impact cardiomyocyte excitation-contraction mechanisms

Finally, to test whether these antibodies could impact cardiomyocyte function, we treated iPSC-CMs with β-AR-specific MI-mAbs. As shown in Figure 4A, MI-mAbs, 2.1, 16.2, 19.2, 79.2, and 116.2 resulted in increased beating frequency. To further investigate whether these mAbs could induce intracellular cAMP signaling downstream of β-AR stimulation, we used HEK cells expressing β1-AR together with a highly sensitive fluorescence resonance energy transfer (FRET)-based cAMP biosensor ^23^. In this biosensor, enhanced YFP and CFP sequences are fused to a cAMP binding domain, and cAMP-induced conformational changes decrease FRET signal, which can be monitored in real time. As expected, the β-AR agonist isoproterenol produced a rapid decrease in FRET, indicating an increase in cytosolic cAMP levels (Fig. 4B). Strikingly, MI-mAbs 2.1, 19.2 and 16.2 induced higher intracellular cAMP levels upon treatment, in line with their β-AR binding specificity, whereas no significant effects were observed in cells treated with isotype control IgG1.

**Fig. 4:**
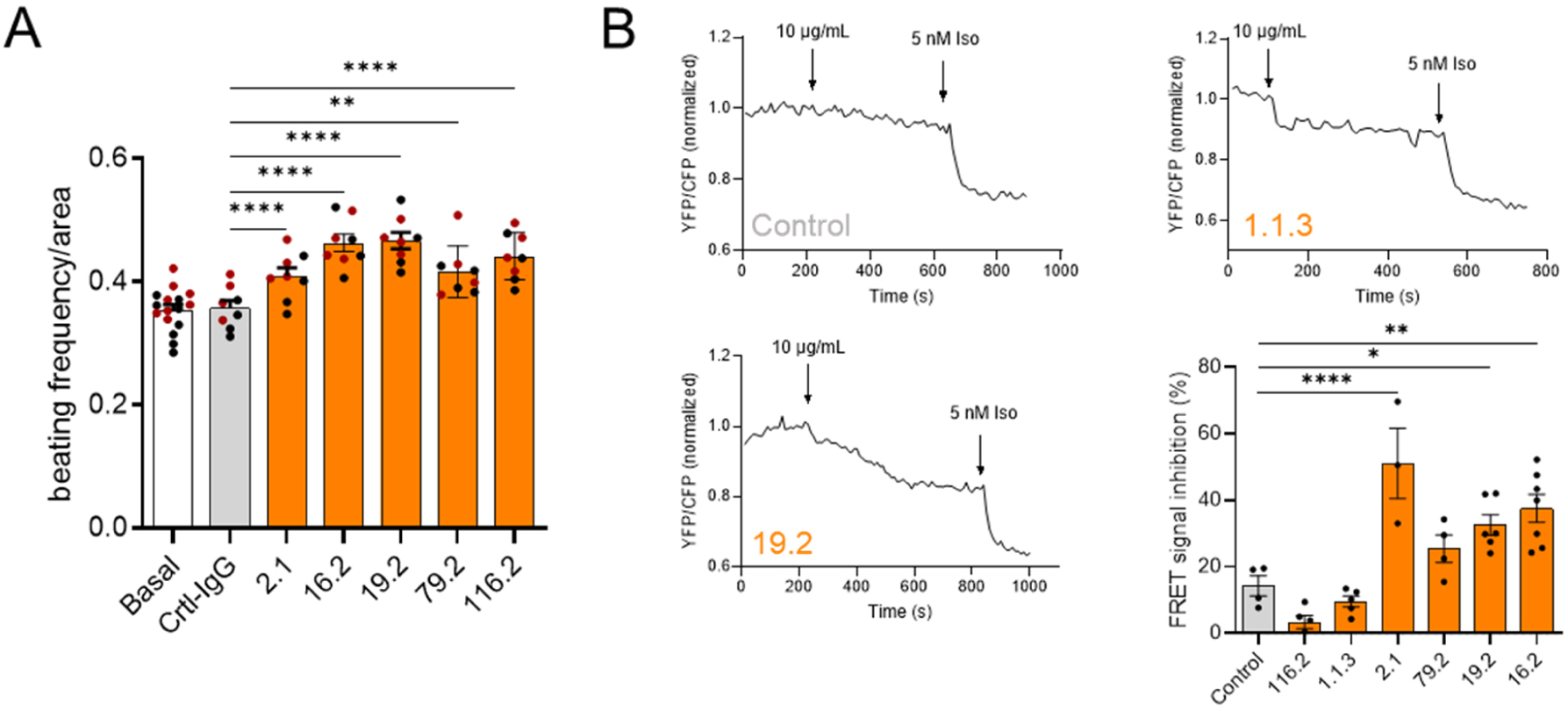
β-AR antibodies impact cardiomyocyte excitation-contraction mechanisms. **(A)** Beating frequency of iPSC-CM is enhanced after treatment with 1 µg/ml of MI-mAbs compared to Ctrl-IgG. **(B)** FRET responses of a HEK cell line expressing the FRET-based cAMP biosensor stimulated first with either MI-mAbs or Ctrl-IgG and subsequently with the β-AR agonist isoproterenol (5 nM Iso). Data are represented as bar graphs with mean ± SEM. *p<0.05, **p<0.01, ***p < 0.001, ****p < 0.0001 = significant differences calculated by 1-way ANOVA with Tukey’s multiple comparison.

Taken together, this scBCRseq approach combined with antibody engineering and epitope mapping reveals that MI triggers B cell responses targeting conserved β-AR extracellular epitopes in MI patients.

## DISCUSSION

The cardiomyocyte necrosis triggered by ischemia exposes cardiac proteins and subcellular components to the immune system within an inflammatory context. This process can ultimately trigger the activation of antigen-specific T and B cells and the production of cardiac plasma antibodies ^17,24^. However, precise mechanistic studies in this field have been impeded by the lack of defined recombinant mAbs. By integrating scBCR repertoire analysis with antibody engineering and iPSC-CM testing, we mapped shifts in BCR repertoires following MI in humans and engineered monoclonal antibodies mimicking those produced in vivo. Our study revealed the induction of anti-CM plasmablast responses in MI patients. Moreover, functional characterization of 17 human mAbs showed that post-MI B cell responses target cardiac receptors and impact cardiomyocyte excitation-contraction mechanisms, demonstrating that the mAbs can serve as precision tools for dissecting patient-specific immune mechanisms in MI and HF.

While antibodies can be useful biomarkers for tissue-specific injury, their pathophysiological relevance in MI and HF remains unclear. Cardiac sarcomere proteins, including myosins and troponins, are abundantly expressed in the heart and therefore frequently targeted by antibodies upon injury ^25,26^. A previous study reported that MI patients with positive serology for troponin-specific antibodies showed significantly reduced ejection fraction ^9^. However, since antibodies cannot access intracellular sarcomere proteins in intact cells, antibodies targeting them are more likely to bear diagnostic rather than therapeutic potential. In contrast, antibodies targeting extracellular epitopes on receptors involved in cardiomyocyte excitation-contraction could provide more compelling mechanistic links to HF pathophysiology. Our study provides an experimental work frame enabling the identification and mechanistic characterization of patient-specific antibodies bearing potential relevance for myocardial disease progression.

Independent studies have reported serum antibodies targeting cardiac receptors, such as muscarinic and adrenergic receptors ^6–8,27^. Specifically, some of those serum reactivities targeting the β1-AR second extracellular loop have been shown to exert agonistic activity ^28,29^. However, the low titers and seroprevalence of these antibodies in MI patients pose significant challenges when studying them using standard serological approaches. Our approach offers a sensitive way to identify β-AR-specific B cells in MI patients and to characterize their molecular antibody characteristics and phenotype. Further, using iPSC-CM to functionally screen for MI-mAbs that impact cardiomyocyte contractility can more efficiently identify patients producing β-AR-specific antibodies with agonistic activity, which may be detrimental in chronic stages. It is also worth noting that the presence of β-AR-specific IgGs in MI patients provides indirect evidence for CD4^+^ T cells targeting the same proteins, since high-affinity antibody production typically requires T cell help ^21,30–32^.

Overall, by integrating BCR repertoire analysis with antibody cloning, cardiomyocyte functional characterization, and epitope mapping, our study identifies antibodies targeting cardiac adrenergic receptors able to impact cardiomyocyte excitation and contraction mechanisms. In addition to providing key mechanistic insights into the pathophysiology of MI, our study provides a translational perspective and recombinant antibody tools to dissect immune mechanisms underlying myocardial diseases on a patient-specific basis.

## MATERIALS AND METHODS

### Methods

#### Human samples

To investigate the presence of autoantibodies in the plasma of patients with myocardial infarction (MI), we conducted the ANTIK-MI (Antikörper-Produktion durch B-Zellen nach Myokardinfarkt) study at the University Hospital Würzburg. Between January and September 2022, 10 patients (9 male, 1 female, 47-71 years) who were diagnosed with acute MI by ECG criteria (STEMI), creatine kinase (CK) levels between 1,000 U/l and 3,000 U/l, and elevated lactate dehydrogenase (LDH) were recruited to the study. All patients in the MI group received cardiac catheter examination and, in most cases, percutaneous coronary intervention. Their blood was collected into CPT heparinized vacutainers within the first 36 hours after hospitalization (baseline), with follow-ups at day 5, after 4 months, and after 12 months. We isolated peripheral blood mononuclear cells (PBMCs) and cryopreserved them in dimethyl sulfoxide (DMSO) for later analysis. Additionally, patients’ plasma was stored at -80°C for further use. Exclusion criteria were active tumor disease, hematologic disease, muscle disease, autoimmune disease, concomitant immunomodulatory medication, and ongoing infection. In addition to a clinical examination, age, sex, cardiovascular risk factors, and laboratory results were logged (Table S1). Samples from age-matched healthy subjects from the population-based STAAB cohort study served as controls ^14^. The study (240/20-am) was approved by the ethics commission at the University Hospital Würzburg. The study met all criteria of the Declaration of Helsinki, and all patients provided informed consent.

#### Cell sorting and single-cell sequencing

Single cells were isolated from PBMCs of four patients at three time points (baseline, day 5, 4 months). Cells were stained with surface antibodies and additionally with anti-CD27, anti-CD38, and anti-CXCR4 Cite-Seq antibodies (BioLegend TotalSeq antibodies, 1:200 dilution). Before sorting, cells were stained with distinct hashtag antibodies for each patient and time point to investigate individual dynamic changes in BCR repertoires. Due to larger sample size, we sequenced two libraries. The first library included the following samples and TotalSeq-C antibodies: P4 baseline (C0251), P4 day 5 (C0252), P4 4 months (C0253), P5 baseline (C0254), P5 day 5 (C0255), P5 4 months (C0256). The second library contained the following samples and TotalSeq-C antibodies: P7 baseline (C0251), P7 day 5 (C0252), P7 4 months (C0253), P9 baseline (C0254), P9 day 5 (C0255), P9 4 months (C0256). Plasmablasts (CD19^+^CD38^+^CD27^+^), memory B cells (CD19^+^CD27^+^CD38^int^), and naive B cells (CD19^+^CD29^-^CD38^+^) were sorted into tubes containing PBS+FCS.

Single cells were partitioned into nanoliter-scale Gel Bead-In-Emulsions (GEMs) using the Chromium™ Controller and processed with Chromium Next GEM Single Cell 5’ v1.1 kits for reverse transcription, cDNA amplification, and library construction, according to the manufacturer’s specifications (10x Genomics, Pleasanton, CA, USA). Amplification and incubation steps were performed using a SimpliAmp Thermal Cycler (Applied Biosystems, Foster City, CA, USA). Libraries were quantified using a Qubit™ 3.0 fluorometer (Thermo Fisher) and quality-checked with a 2100 Bioanalyzer using High Sensitivity DNA kits (Agilent, Santa Clara, CA, USA).

Libraries were then pooled and sequenced on the NovaSeq 6000 platform (S2 Cartridge, Illumina, San Diego, CA, USA) in paired-end mode to achieve at least 70,000 reads per single cell for gene expression and 7,000 reads for the B-cell receptor repertoire and hashtags. The Cell Ranger Software Suite (10x Genomics) was used for sequence alignment, barcode processing, and sample demultiplexing. Data analysis involved the Seurat R package v3.1.420 for filtering low-quality cell data, normalizing gene expression, and clustering. For B-cell receptor repertoire analysis, the scRepertoire R package (https://f1000research.com/articles/9-47/v1) was utilized.

#### In silico analyses of B cell receptor repertoire data

The V(D)J sequences for paired Ig light and heavy chains from individual cells were combined using combineBCR() and then added to the Seurat object using the combineExpression() function from scRepertoire R package ^33^. For mutation frequency calculation and BCR lineage tree construction, the Immcantation framework was utilized. Briefly, the Change-O and IgBlast packages were employed to generate Adaptive Immune Receptor Repertoire (AIRR)-compliant V, D, and J rearrangement data from 10x V(D)J BCR FASTA files. Cells with multiple functionally rearranged Ig heavy chain transcripts or no Ig heavy chain transcripts were excluded from the analysis, and cellular barcode identities were matched between gene expression and BCR data. Unmatched cells were removed. VDJ rearrangements of the heavy chains were analyzed using the shazam R package to identify clonally related lineages based on the nearest-neighbor distance distribution of CDR3 junction sequences. A Gamma/Gaussian Mixture Model was fitted to establish the threshold for clonal assignment. Subsequently, the scoper R package was employed to cluster sequences with shared V gene, J gene, and junction length from merged heavy and light chains into clonal lineages.

Clonal lineage analysis was performed separately for each individual donor. The unmutated common ancestor for each clonal lineage was reconstructed using the Dowser and shazam packages, and the median mutation frequency of clones was calculated. Finally, standard maximum likelihood trees were built with Dowser to generate B cell phylogenetic trees.

#### Directional cloning and production of monoclonal antibodies

Sequences of Ig heavy and light chain variable regions were purchased from IDT as double-stranded DNA fragments (gBlocks™). DNA fragments were dissolved and amplified in line with the manufacturers’ instructions. Expression vectors AbVec2.0-IGHG1 (Addgene ID 80795) and AbVec1.1-IGKC (Addgene ID 80796) encoding for the human constant regions were purchased from Addgene. Restriction digestions were performed using the following restriction enzymes from NEB: AgeI-HF (IGHG1, IGKC), SalI-HF (IGHG1), and BsiWI-HF (IGKC) ^16^. Correct cloning was confirmed after Sanger sequencing. FreeStyle™ 293-F-cells were co-transfected with IgH and IgK chain plasmids. Six days after transfections, supernatants were harvested by two consecutive centrifugations at 4,000 rpm for 30 min. Supernatants were incubated with Protein G beads (1:1,000) overnight at 4°C under rotation. The next day, recombinant monoclonal antibodies were purified from the supernatant by using chromatography columns (Biorad) equilibrated with PBS (protocol adapted from Tiller, T. et al ^34^). After two consecutive washes with 1 ml PBS each, antibodies were eluted in three fractions with 0.1 M glycine (pH = 3) and collected in falcons containing 1 M Tris with 0.5% sodium azide (pH = 8). Finally, IgG concentrations were measured by human IgG ELISA.

#### Immunofluorescence

For immunofluorescence analyses of human-derived iPSC-CMs, cells were fixed with 4% FA. The cells were permeabilized with 0.1% Triton X 100 in PBS and blocking was performed in 0.1% Triton X 100 in PBS with 1% BSA. After fixation, cells were incubated with either 10 µg/ml of each recombinant monoclonal antibody or a 10-fold dilution of plasma in blocking buffer. Additionally, cells were counter-stained with either mouse anti-alpha actinin (1:500) or rabbit anti-TTN-9 (1:300). The following fluorophore-conjugated secondary antibodies were used: goat anti-human IgG Alexa Fluor 555 (Invitrogen, 1:200), goat anti-mouse IgG Alexa Fluor 488 (Invitrogen, 1:200), and donkey anti-rabbit IgG Alexa 647 (Invitrogen, 1:500). During the final washing step, cells were incubated with Hoechst 33342 (B2261, Sigma-Aldrich, 10 mg/ml) using a 1:3,000 dilution in PBS for 5 min at room temperature (RT). The cells were imaged using a laser scanning confocal microscope (LSM 980 Airyscan 2, Zeiss, Oberkochen, Germany) and a 63x objective (oil immersion). The acquired multichannel images were deconvolved with Huygens Professional (Scientific Volume Imaging, Hilversum, Netherlands) (raw images available upon request). The processed images were loaded into the Fiji distribution of ImageJ for contrast adjustment and saved in png-format for visualization ^35^.

#### ELISAs

Total IgG concentrations were determined using a sandwich enzyme-linked immunosorbent assay (ELISA, Invitrogen), following the manufacturer’s instructions. To summarize, plates were coated with anti-human IgG coating antibodies (1:250), blocked with blocking buffer (1:10 dilution of assay buffer A (20X) in deionized water), and incubated with purified IgG samples (10-fold serial dilutions with assay buffer A ranging from 1:100 to 1:100,000).

Wells were incubated with anti-human IgG-HRP detection antibodies (1:250), followed by addition of substrate solution. 1 M H_3_PO_4_ was then added to stop the reaction. Optical density (OD) was measured at 450 nm with a reference wavelength of 570 nm.

Further, to investigate mAb binding to ß-ARs, we performed indirect ELISAs by coating 5 µg/well with an extracellular peptide pool comprising either peptides of ADRB1 (MGAGVLVLGASEPGNLSSAAPLPDGAATAARLLVPASPPASLLPPASESPEPLSQ, WGRWEYGSFFCE, HWWRAESDEARRCYNDPKCCDFVTNR) or peptides of ADRB2 (MGQPGNGSAFLLAPNGSHAPDHDVTQERDEVWVV, MKMWTFGNFWC, RATHQEAINCYANETCCDFFTN) diluted in carbonate buffer (pH 9.5). Wells were blocked using ELISA Assay Diluent (1X in PBS), which was also used to dilute monoclonal antibodies. Antibodies (10 μg/ml, 100 μl/well) were applied in duplicates followed by incubation with HRP-conjugated anti-human IgG antibody (Thermo Fisher, 1:2,000 in blocking solution). HRP substrate (BioLegend) was added, the reaction was stopped with 2 N H₂SO₄, and absorbance was measured at 450 nm with 570 nm as reference. Background-subtracted means of duplicates were used for analysis.

#### Differentiation of iPSCs into iPSC-CMs and quality control

Induced pluripotent stem cells (iPSCs) were generated from healthy donors without any known cardiovascular disease ^36^. All subjects gave written informed consent regarding tissue donation. iPSC generation was approved by the local ethics committee of the University of Goettingen (10/9/15), with the donors’ written consent. The iPSC lines were cultured feeder-free and adherend on cell culture dishes in chemically defined medium E8 (Life Technologies).

Cardiac differentiation of iPSCs to ventricular iPSC-cardiomyocytes (iPSC-CMs) was performed by sequentially targeting the Wnt pathway as described previously ^37^. Briefly, undifferentiated iPSCs were cultured as a monolayer on Geltrex-coated 12-well dishes. iPSCs differentiation was initiated by changing the medium to RPMI 1640 (Thermo Fisher) supplemented with 0.02% L-ascorbic acid 2-phosphate (Sigma-Aldrich) and 0.05% albumin (Sigma-Aldrich) including the GSK3 inhibitor CHIR99021 (4 μmol/L; Millipore) at 85% to 90% confluence (d0). After 48 hours, medium was changed to fresh medium supplemented with 5 μmol/L of IWP2 (Wnt signaling inhibitor; Millipore) for 2 days. From day 10 on, cells were cultured in cardio culture medium (RPMI 1640) supplemented with 2 mmol/L L-glutamine and 2% B27 with insulin (Life Technologies). The medium was changed every 2 to 3 days. iPSC-CMs were purified by metabolic selection. iPSC-CM purity was determined by flow analysis (>80% cardiac troponin T positive) and qPCR analysis for cardiac ventricular sub-type markers (*MLC2v* vs. *NR2F2*). Experiments were carried out during days 60-90 after differentiation initiation. Cells were incubated with respective IgGs (1 µg/ml) acutely (30 min-1h) or chronically (3-5 days).

#### Beating frequency of iPSC-CMs

Ventricular iPSC-CMs between days 60 and 90 were digested, plated on Geltrex-coated 6-well plates, and incubated in cardio culture medium for 1 week before use. For chronic treatment, iPSC-CMs were incubated with respective IgGs (1 µg/ml) in cardio culture medium for 5 days. Fresh medium with/without antibodies was given every 2-3 days and 24 hours before starting the measurements. For acute treatment, medium was changed 24 hours before starting the measurements and iPSC-CMs were incubated with respective IgGs (1 µg/ml) in cardio culture medium for 30 min. iPSC-CMs were adjusted to RT for 10 minutes before beating frequency measurements. Beating frequency of iPSC-CM was measured using the Ion optics/ Ion wizard system.

#### Viability test (Resazurin assay)

Ventricular iPSC-CMs between days 60 and 90 were plated onto a Geltrex-coated 96-well plate and incubated in cardio culture medium for 1 week before use. These cells were incubated with respective antibodies (MI-mAbs, Ctrl-IgG; 1 µg/ml) for 30 min for acute and 3 days for chronic treatment. Cells were incubated with IgGs for 1 hour for acute treatment before the Resazurin assay was performed. To establish a zero-viability control, we used 10% DMSO, which induces cell membrane permeabilization. This assay reveals the antibody treatments’ effects on iPSC-CM viability. Specifically, the assay can estimate cell viability by measuring the metabolic activity of viable cells capable of converting blue resazurin into pink resorufin ^38^. Resazurin (Cayman Chemical Co.) was prepared as a stock solution of 14 mM in PBS and used at a final concentration of 70 μM. The absorbance of resorufin, proportional to the number of viable cells, was detected by a standard spectrophotometer at 540 nm and 590 nm.

#### Förster resonance energy transfer (FRET) live-cell measurements of cAMP

Human embryonic kidney HEK293 cells stably expressing human β1-adrenergic receptor (β1-AR) and the cytosolic Epac1-based fluorescent cAMP sensor (Epac1-camps) were used for cAMP measurements. FRET measurements were performed using an inverted fluorescent microscope (Leica DMI 3000B) with an oil-immersion 63x/1.4 objective. Cyan fluorescence protein (CFP) was excited at 440 nm using a pE-100 CoolLED device. The emitted light was split into donor and acceptor channels by DV2 Dual View beam splitter (Photometrics; Cube 05-EM, 505 dcxr, D480/30, D535/40) and recorded on a CMOS (OptiMOS, QImaging) camera chip. During the measurements, cells were maintained in FRET buffer (144 mM NaCl, 5.4 mM KCl, 1 mM CaCl_2_, 1 mM MgCl_2_, 10 mM HEPES, pH 7.3). The antibody preparations were diluted in FRET buffer to a final concentration of 10 µg/mL. To determine the maximal cAMP response, 5 nM Isoproterenol was added in the end of each measurement. Images were taken every 10 s using MicroManager 1.4 software. Data analysis was performed in ImageJ, followed by a raw data correction for a spectral bleed-through factor carried out in Microsoft Excel.

#### Peptide microarray

The custom peptide microarray included a list of 42 cardiac membrane proteins, containing adrenergic and muscarinic receptors, as well as ion channels. These proteins were analyzed computationally to identify potential extracellular continuous and discontinuous (conformational) epitopes. The final library consisted of 6,543 unique 15-mer peptides, which were displayed in duplicates in a multi-well layout. Microarray fabrication, incubation with analytes and data processing was performed by Axxelera (Karlsruhe, Germany). Peptides with normalized signal values greater than 3 were considered true positive hits, whereas values below -3 were treated as potential artifacts or negative outliers. To identify robust antibody–epitope interactions, peptides with normalized signals above 3 were grouped by their corresponding protein. High-confidence continuous epitope regions were defined as overlapping peptide fragments with start positions shift by exactly three amino acids. For the identification of potential discontinuous (structural) epitopes, peptides with normalized signals above 3 were evaluated. To enhance reliability and minimize background noise, only those discontinuous peptides that were detected more than once in the dataset were included in the final analysis.

## Data Availability

All data produced in the present study are available upon reasonable request to the authors

## List of non-standard abbreviations

β1-AR: β1-adrenergic receptor
β2-AR: β2-adrenergic receptor
BCR: B cell receptor
HF: heart failure
iPSC-CM: induced pluripotent stem cell-derived cardiomyocyte
LN: lymph node
MI: myocardial infarction
MI-mAb: myocardial infarction induced monoclonal antibodies
PB: plasmablast
Ig, IgA, IgG, IgM: immunoglobulin
scRNA/BCRseq: single-cell RNA / BCR sequencing

## Acknowledgments

The authors thank Elena Vogel, Yvonne Metz, and Bianca Klüpfel for their skillful technical assistance. We thank the Core Unit SysMed at the University of Würzburg for excellent technical support, RNA-seq data generation, and analysis.

## Funding

German Research foundation (DFG), Collaborative Research Centre 1525 ‘Cardio-immune interfaces’, (grant number 453989101 - project B6 to GCR and KGH)

German Research foundation (DFG), the Heisenberg Program, grant 517001338 (GCR)

German Research foundation (DFG), grant INST 93/1022-1 (KGH)

German Research foundation (DFG), grant 471241922 (KSB)

German Research foundation (DFG), Collaborative Research Centre 1213 ‘Pulmonary Hypertension and Cor Pulmonale (KSB)

German Federal Ministry of Education and Research, BMBF 01EO1004 and 01EO1504 (SS, PH)

Interdisciplinary Center for Clinical Research of the University Hospital Würzburg, grant IZKF-Z12 and IZKF-Z15 (KGH)

## Author contributions

Conceptualization: JS, KGH, GCR; Methodology: JS, AA, DA, HW, VN, ES, CC, SS, PH, CM, MH, KGH, KSB, GCR; Investigation: JS, DA, AA, DB, SJ, LSZ, EK, MH, MB, TK; Visualization: JS, AA, SJ, MB, KSB, GCR; Funding acquisition: KGH, KSB, GCR, SF; Project administration: KGH, GCR; Supervision: KGH, KSB, GCR; Writing – original draft: JS, GCR; Writing – review & editing: all.

## Data and materials availability

Anonymized patient sequencing data will be made available upon reasonable request.

