## Supplemental Data for "Dissecting antibody responses to cardiac receptors in patients with myocardial infarction"

**Affiliations:**

Prof. Dr. Gustavo Ramos

Immunocardiology Laboratory

University Hospital Würzburg, Comprehensive Heart Failure Center

Am Schwarzenberg 15, Haus A15

D-97078 Würzburg, Germany

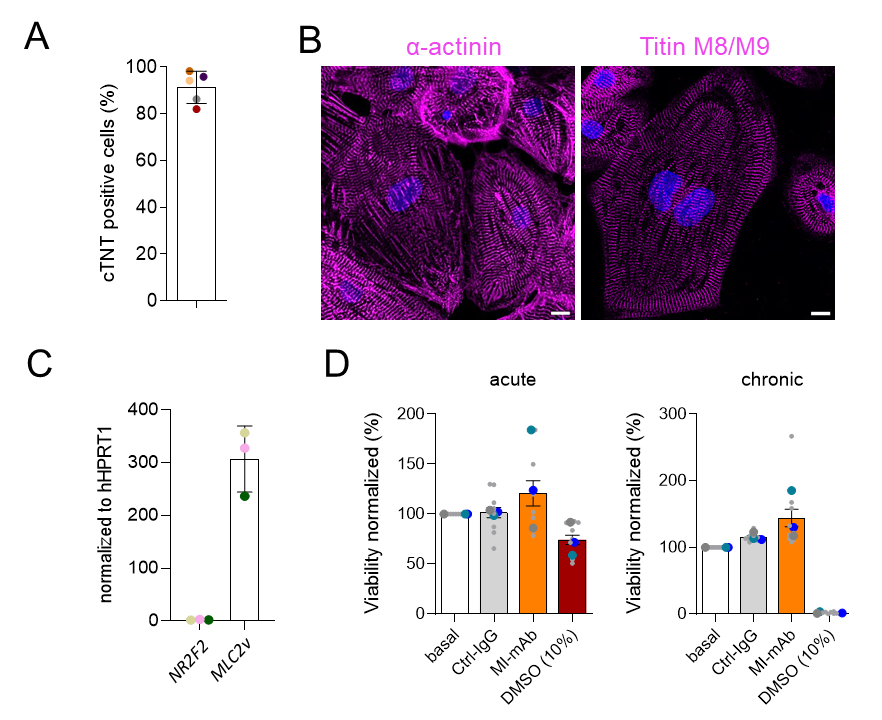

Fig. S1: Quality control of iPSC-CMs. (A) The purity of the iPSC-CMs was determined by flow cytometry for cardiac troponin T. (B) Staining of ventricular iPSC-CMs with α-actinin and titin M8/M9. magenta: α-actinin/titin M8/M9; blue: Hoechst. Scale bar: 10 µm. (C) qPCR analysis for cardiac subtype-specific markers (*NR2F2*: atrial; *MLC2v*: ventricular). Each dot represents one cardiac differentiation. (D) The viability of the cells treated with Ctrl-IgG and MI-mAbs for 30 min (acute) and 4 days (chronic) was determined by the Resazurin assay. DMSO-treated cells served as a positive control for the viability assessment (not vehicle). Each big dot represents the mean of one cardiac differentiation. Small dots represent different technical replicates. (DMSO = dimethyl sulfoxide).

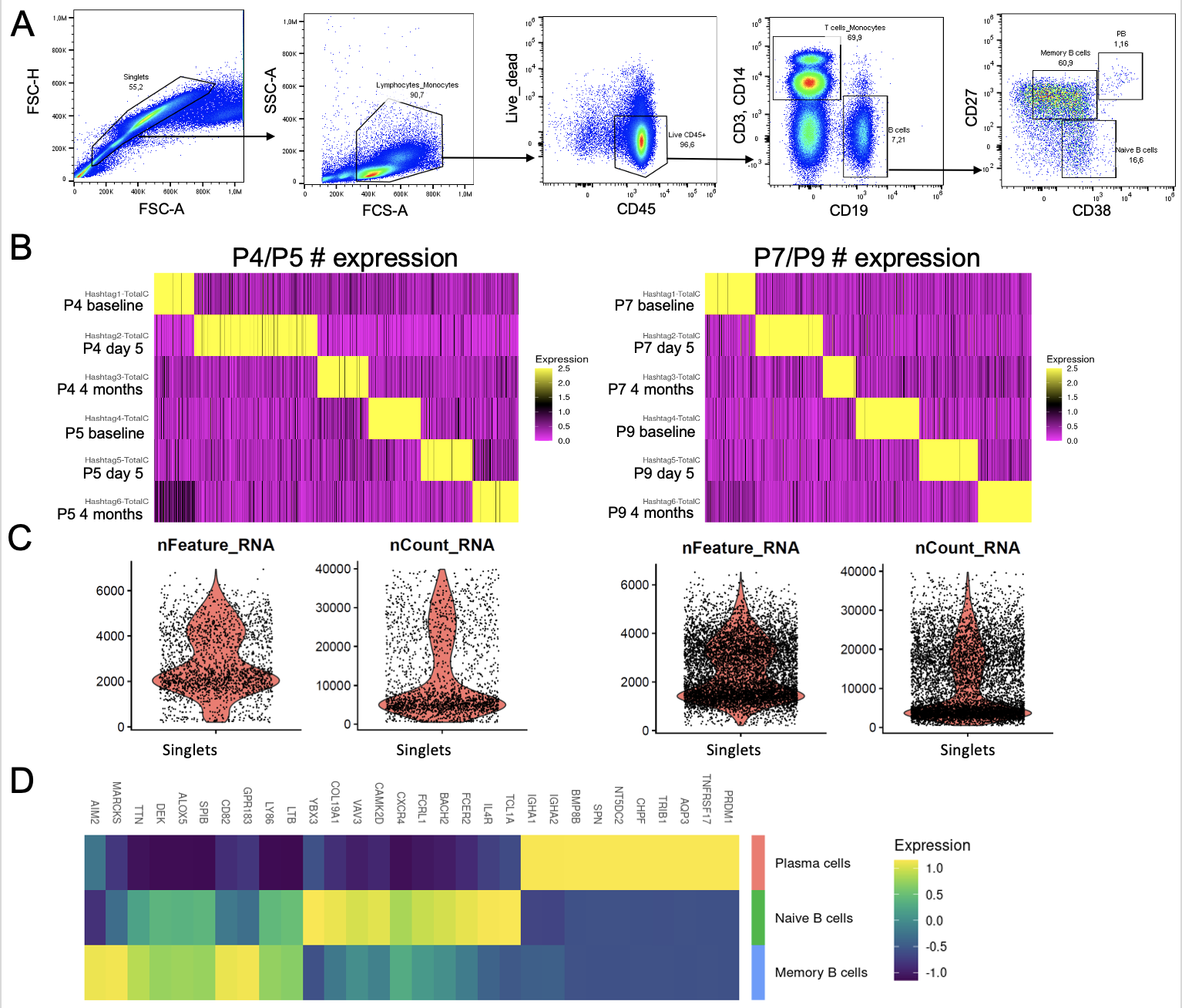

Fig. S2: Expression of cluster-defining transcripts in MI patients. (A) Gating strategy for the flow cytometric isolation of plasmablasts, memory and naive B cells. (B) Heatmap showing the hashtag strategy for demultiplexing B cells from different time points of patients (P) 4 and 5 (left) and patients 7 and 9 (right). (C) Violin plots showing the number of transcripts and RNA count in all B cells of the scRNA objects. (D) Heatmap showing 3 clusters that were identified by top 10 most expressed transcripts.

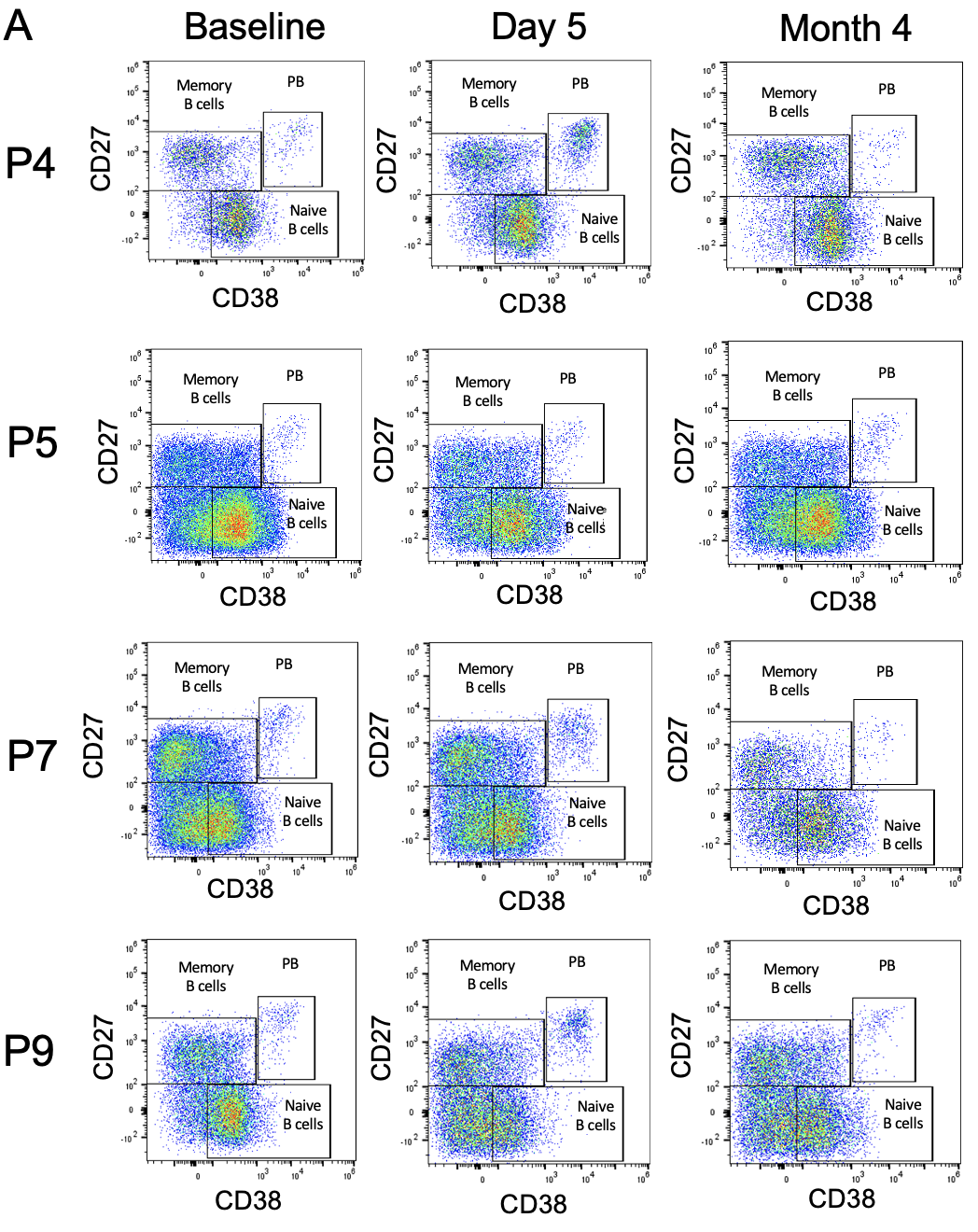

**Fig. S3: Characterization of B cell subsets in MI patients.** Flow cytometry plots showing dynamics in plasmablasts (CD27^high^CD38^high^), memory B cells (CD27^+^CD38^-/+^), and naïve B cells (CD27^-^CD38^int^). Parent gate: Singlets, live, CD45^+^ CD3^-^ CD14^-^ CD19^+^.

Table S1: Characteristics of MI patients included in ANTIK study.

| **Age** | 58 ± 10 |
| --- | --- |
| **Sex** | 9/10 male |
| **MI type** | STEMI (10/10) |
| **Risk factors** | ≥ 1 risk factor: 9/10 |
| **Tabagism** | 8/10 |
| **Adiposity** | 2/10 |
| **Dyslipidemy** | 2/10 |
| **Diabetes Mellitus** | 2/10 |
| **Family History** | 1/10 |
| **Arterial Hypertony** | 7/10 |
| **CK max. (U/l)** | 2,171 ± 1,126 |
| **Troponin (pg/ml)** | 3,267 ± 3,216 |
| Median ± interquartile range | |

Table S2: Overview and characteristics of MI-mAbs.

| MI-mAb | *IGHC* | *IGHV* | *IGHD* | *IGHJ* | Mutation frequency^(1)^ | Clone size^(2)^ |
| --- | --- | --- | --- | --- | --- | --- |
| **1.1.1**  **1.1.2**  **1.1.3** | *IGHG2* | *IGHV3-23*01* | *IGHD1-1*01* | *IGHJ5*02* | Medium  High High | Medium |
| **2.1** | *IGHG2* | *IGHV3-74*01* | *IGHD2-15*01* | *IGHJ3*02* | Medium | Small |
| **4.1** | *IGHG2* | *IGHV1-2*02* | *IGHD3-9*01* | *IGHJ3*01* | High | Small |
| **21.1** | *IGHG1* | *IGHV1-8*01* | *IGHD3-3*01* | *IGHJ5*02* | Large | Small |
| **43.2** | *IGHG2* | *IGHV3-23*01* | *IGHD5-24*01* | *IGHJ4*02* | Medium | Small |
| **23.2**  **113.2** | *IGHG3* | *IGHV1-69*04* | *IGHD3-10*01* | *IGHJ6*02* | High  High | Small |
| **30.2** | *IGHM* | *IGHV3-23*01* | *IGHD2-8*02* | *IGHJ3*02* | Small | Medium |
| **16.2** | *IGHG2* | *IGHV3-7*05* | *IGHD6-13*01* | *IGHJ4*02* | Small | Medium |
| **19.2** | *IGHG1* | *IGHV4-34*01* | *IGHD6-13*01* | *IGHJ4*02* | Small | Medium |
| **79.2** | *IGHG1* | *IGHV4-39*07* | *IGHD2-15*01* | *IGHJ6*02* | Small | Small |
| **116.2** | *IGHG3* | *IGHV4-34*01* | *IGHD4-23*01* | *IGHJ3*02* | Medium | Small |
| **1.2** | *IGHG2* | *IGHV3-30*03* | *IGHD2-2*02* | *IGHJ6*02* | Medium | Large |
| **3.2** | *IGHA1* | *IGHV4-39* | *IGHD5-12*01* | *IGHJ6*02* | N/A | Small |
| **4.2** | *IGHG2* | *IGHV3-30*18* | *IGHD4-11*01* | *IGHJ6*02* | High | Large |

^(1)^ Small (0.00 < X <= 0.05); Medium (0.05 < X <= 0.10); Large (0.10 < X <= 0.15)

^(2)^ Small (1e-04 < X <= 0.001); Medium (0.001 < X <= 0.01); Large (0.01 < X <= 0.1)

Table S3: List of proteins used in the peptide array.

**Protein name**

5-hydroxytryptamine receptor 4

Acetylcholine receptor subunit epsilon

Alpha-1A adrenergic receptor

A-type voltage-gated potassium channel KCND1

A-type voltage-gated potassium channel KCND2

A-type voltage-gated potassium channel KCND3

Beta-1 adrenergic receptor

Beta-2 adrenergic receptor

Muscarinic acetylcholine receptor M2

Potassium voltage-gated channel subfamily A member 1

Potassium voltage-gated channel subfamily A member 4

Potassium voltage-gated channel subfamily A member 5

Potassium voltage-gated channel subfamily A member 6

Potassium voltage-gated channel subfamily A member 7

Potassium voltage-gated channel subfamily B member 1

Potassium voltage-gated channel subfamily E regulatory beta subunit 5

Sodium channel protein type 1 subunit alpha

Sodium channel protein type 10 subunit alpha

Sodium channel protein type 2 subunit alpha

Sodium channel protein type 3 subunit alpha

Sodium channel protein type 4 subunit alpha

Sodium channel protein type 5 subunit alpha

Sodium channel protein type 8 subunit alpha

Sodium channel regulatory subunit beta-1

Sodium channel regulatory subunit beta-2

Sodium channel regulatory subunit beta-3

Sodium channel regulatory subunit beta-4

Sodium/calcium exchanger 1

Sodium/potassium-transporting ATPase subunit alpha-2

Type-1 angiotensin II receptor

Type-2 angiotensin II receptor

Voltage-dependent calcium channel subunit alpha-2/delta-1

Voltage-dependent L-type calcium channel subunit alpha-1C

Voltage-dependent L-type calcium channel subunit alpha-1S

Voltage-dependent P/Q-type calcium channel subunit alpha-1A

Voltage-dependent T-type calcium channel subunit alpha-1G

Voltage-dependent T-type calcium channel subunit alpha-1H

Voltage-gated delayed rectifier potassium channel KCNH4

Voltage-gated inwardly rectifying potassium channel KCNH2

Voltage-gated inwardly rectifying potassium channel KCNH3

Voltage-gated potassium channel KCNC3

Voltage-gated potassium channel regulatory subunit KCNG2
